# COVID-Q: validation of the first COVID-19 questionnaire based on patient-rated symptom gravity

**DOI:** 10.1101/2021.07.24.21261065

**Authors:** G Spinato, C Fabbris, F Conte, A Menegaldo, L Franz, P Gaudioso, F Cinetto, C Agostini, G Costantini, P Boscolo-Rizzo

## Abstract

**Objectives:** The aim of the present study was to develop and validate the CoronaVirus Disease – 2019 (COVID-19) Questionnaire (COVID-Q), a novel symptom questionnaire specific for COVID-19 patients, to provide a comprehensive evaluation which may be helpful for physicians.. A secondary goal of the present study was to evaluate the performance of the COVID-Q in identifying subjects at higher risk of being tested positive for COVID-19.

**Material and methods:** Consecutive non-hospitalized adults who underwent nasopharyngeal and throat swab for severe acute respiratory syndrome coronavirus 2 (SARS-CoV-2) detection at Treviso Hospital in March 2020, were enrolled. Subjects were divided into positive (cases) and negative (controls) in equal number. All of them gave consent and answered the COVID-Q. Patients not able to answer the COVID-Q due to clinical conditions were excluded.

Parallel Analysis and Principal Component Analysis were used to identify clusters of items measuring the same dimension. The Item Response Theory (IRT)-based analyses evaluated the functioning of item categories, the presence of clusters of local dependence among items, item fit within the model and model fit to the data.

**Results:** Answers obtained from 230 COVID-19 cases (113 males, and 117 females; mean age 55 years, range 20-99 years) and 230 controls (61 males, and 169 females; mean age 46 years, range 21-89) were analyzed. Parallel analysis led to the extraction of six components, which corresponded to as many clinical presentation patterns: asthenia, influenza-like symptoms, ear and nose symptoms, breathing issues, throat symptoms, and anosmia/ageusia. The final IRT models retained 27 items as significant for symptom assessment. The total score on the questionnaire was significantly associated with positivity to the molecular SARS-CoV-2 test: subjects with multiple symptoms were significantly more likely to be affected by COVID-19 (p < .001). Older age and male gender also represented risk factors. Presence of breathing issues and anosmia/ageusia were significantly related to positivity to SARS-CoV-2 (p < 0.001). None of the examined comorbidities had a significant association with COVID-19 diagnosis.

**Conclusion:** According to the analyses, COVID-Q could be validated since the aspects it evaluated were overall significantly related to SARS-CoV-2 infection. The application of the novel COVID-Q to everyday clinical practice may help identifying subjects who are likely to be affected by COVID-19 and address them to a nasopharyngeal swab in order to achieve an early diagnosis.

**What is already known about this topic?:** COVID-19 symptoms are widely known. Lots of studies have been published regarding self-administered questionnaires in order to characterize and know as much as possible regarding this disease. By the way, no specific questionnaires have been validated, yet, and there is no consensus regarding this topic.

**What does this article add?:** This paper shows the COVID-Q, a novel symptom questionnaire specific for COVID-19 patients. The aim is to provide a comprehensive evaluation that may be helpful to clinicians in order to suspect SARS-CoV-2 infection or not.

## INTRODUCTION

COVID-19 is an emerging infectious disease due to SARS-CoV-2, whose worldwide outbreak led, during late 2019 and during 2020, to the current pandemic condition^1-3^.

The clinical presentation of COVID-19 ranges from the complete absence of symptoms to a severe acute respiratory distress syndrome, potentially leading to death^1,4,5^. In symptomatic patients, the most common presentation includes fever, myalgia and fatigue, associated with upper and lower respiratory tract involvement, resulting in nasal congestion, sore throat, anosmia, ageusia, dry cough, and dyspnea^4^. In addition to these respiratory symptoms, gastrointestinal^6^, cardiovascular^7^ and neurological^8-9^ manifestations have also been described.

Among the upper airways symptoms, the new onset of an altered sense of smell or taste has been regarded with particular interest as clinical hallmark of early SARS-CoV-2 infection, potentially able to discriminate subjects with COVID-19 from those with other types of acute respiratory tract infections^10-17^.

Given the critical importance of a comprehensive assessment of the severity of COVID-19 clinical presentation, risk factors and comorbidities useful to characterize patients who will develop a severe COVID-19 have been discussed.^18^ On the other hand, currently there is a lack of validated and standardized means to evaluate symptoms.

Clinical evaluation models based on patient-reported outcomes (PROs) have been regarded with increasing interest, in particular in the field of pneumology, to evaluate conditions such as asthma, pneumonia, and chronic obstructive pulmonary disease (COPD)^19-22^.

To date, only a PRO questionnaire, with adequate content validity and psychometric properties, has been developed and validated to measure the severity, functional impact and response to therapy of acute respiratory tract infections in adults^23^. Such reporting tool, the Acute Respiratory Tract Infections Questionnaire (ARTIQ)^23^, resulted to be effective in discriminating between participants with and without acute respiratory tract infections.

To the best of our knowledge, no standardized tool has been proposed yet for a uniform and comprehensive evaluation and reporting of symptoms in COVID-19 patients.

The present study aimed to develop and validate the COVID-Q, a novel questionnaire specific for COVID-19 patients in order to provide a comprehensive and standard clinical evaluation. A secondary goal of the present study was to evaluate the performance of the questionnaire in identifying subjects at higher risk of being tested positive for COVID-19.

## MATERIALS AND METHODS

This study was approved by the Ethical Committees of the Academic Hospital of Treviso (Italy) and the Hospital of Belluno (Italy), and it was conducted in accordance with the principles of the Helsinki Declaration. Data were examined in compliance with Italian privacy and sensitive data laws. All participants gave their consent to be enrolled in this investigation and to the treatment of their personal data for scientific purposes. The study was conducted in the Hospital of Treviso.

### Questionnaire construction

All patients both in the case and the control group answered the COVID-Q.The COVID-Q included items focused on general, ENT, respiratory and gastrointestinal symptoms, as well as psychological symptoms, drug assumption and changes in daily activity. Such items in part belonged to the ARTIQ^23^ and the SNOT-22^244^ questionnaires, and the remaining were developed *ex novo* by us to better characterize symptoms and clinical conditions specific of COVID-19, basing on the data available from the Atlanta CDC guidelines^25^. All items based on the ARTIQ were scored on a three-point scale (0 = none, 1 = a little, 2 = a lot) in line with the ARTIQ itself, while the questions focused on anosmia and ageusia, were scored on a scale from 0 (none) to 5 (complete loss of smell or taste), being derived from the SNOT-22. An additional section of the questionnaire focused on demographic data, including age, sex, smoking status (0 = never, 1 = former smoker, 2 = current smoker) and comorbidities (0 = absent, 1 = present).

All subjects in both groups answered the questionnaire via a telephone interview, which took place within 19 days from the first diagnosis in cases and within 43 days from the last negative test in controls.***Participants*** Consecutive subjects who underwent nasopharyngeal and throat swabs for SARS-CoV-2 detection were included in the study. People were tested in the hospital of Treviso between March 4^th^ and 24^th^, 2020. Tests of SARS-CoV-2 RNA by polymerase chain reaction on nasopharyngeal and throat swabs were performed according to the World Health Organization recommendation^25^. Reasons for being tested were symptoms, close contact with affected individuals or working in public health services. Non-hospitalized adults (≥18 years) were considered eligible for the study. The whole sample was split into two arms: “cases” who tested positive for SARS-CoV-2 infection and “controls” who tested negative.

The inclusion criteria were the consent to take part to the study and positive test for COVID-19 (for inclusion in the cases group). The main exclusion criteria were clinical conditions leading to the inability to answer the COVID-Q (severe respiratory syndrome, hospitalization in the Intensive Care Unit, cognitive impairment). All subjects in both groups answered the questionnaire via a telephone interview.

### Statistical Analyses

The statistical analyses were performed only on 41 of the items from the ARTIQ and SNOT-22, since the remaining regarded self-administrated drugs and social activities, which were not taken into account due to the peculiar circumstances of data collection. Indeed, participants were tested during the first lockdown phase in Italy, when social and work activities had been heavily restricted and people were advised not to take any medication without consulting with their physician.

First, Parallel Analysis and Principal Component Analysis (PCA) were used to identify clusters of items measuring the same dimension. Given the ordinal nature of the data in question, the analyses were performed on the polychoric correlation matrix.

IRT models for polytomous items were fit on each of the components identified by the PCA. We first compared the fit of a freely estimated Generalized Partial Credit Model (GPCM)^27^ to a common-slope GPCM, in which the slopes (i.e., discrimination parameters) of all items are constrained to be equal. The IRT-based analyses evaluated the functioning of item categories, the presence of clusters of local dependence among items, item fit within the model and model fit to the data. Items were selected through an iterative procedure. Items for which the three response categories did not function as intended were recoded into two categories: 0 = absence of symptom, 1 = presence of symptom. Local dependence was measured by the Q3 index^28^: following Marais’s^29^ indications, residual correlations between items were compared to the average residual correlation within the dataset, and the greatest deviances were taken to suggest possible item redundancy or underlying dimensions. Item fit was based on the scaled version of the chi square statistic χ^2^*^30^: p-values smaller than .05 were considered as indicating significant misfit and led to the exclusion of the item. Finally, omnibus fit was evaluated by the M2* limited-information test^31^ and was considered acceptable when p values were above the .05 threshold. The items retained in the IRT models were then used to produce scale and total scores, computed as raw sums, and represented the final validated version of the questionnaire.

We estimated the extent to which self-reported symptoms could help identify COVID-19 by means of two different logistic regression models. In both models, the diagnosis (1 = positive, 0 = negative) was the dependent variable, and age, sex, smoking status, and presence of comorbidities were used as covariates. The predictors of interest were the total questionnaire score in the first model, and individual scale scores in the second model. By design, affected individuals were over-represented in our sample compared to the population: this was taken into account by applying the appropriate correction for rare events^32^. Further examinations were performed according to the obtained results to refine items by using the DETECT method in package sirt^33,34^. Finally, we identified questionnaire scores associated with specific risk thresholds for COVID-19.

## RESULTS

### Sample characteristics

A sample of 460 participants (176 males and 286 females; mean age = 51 years, range 20-99 years) was recruited. Among participants there were 230 cases (113 males, and 117 females; mean age 55 years, range 20-99 years) and 230 controls (61 males, and 169 females; mean age 46 years, range 21-89). The telephone interview took place within 19 days from the first diagnosis in cases and within 43 days from the last negative test in controls. The latter time frame resulted to be reliable because, being the controls negative, there was no evolution in their clinical condition, which was comparable with the moment of the first swab.

The prevalence of smoke and comorbidities in cases and controls is shown in **Supplementary Table S1**. An independent sample t-test revealed that the average number of comorbidities (as considered as present or not present) per person was significantly higher among cases than controls (t(11) = 2.263, p = .045). In particular, Fisher’s exact tests showed that hypertension, diabetes, and cardiovascular diseases were significantly more frequent among cases, compared with controls (p < .001, p = .001 and p = .030, respectively). No significant differences in the distribution of the remaining comorbidities or in the proportion of smokers emerged between the two groups. Liver diseases were excluded from all analyses, having been observed in only one case.

### Principal component analysis

Parallel analysis led to the extraction of five components, which corresponded to as many clinical presentation patterns: asthenia (including fatigue, sleep disorders and depression mood), influenza-like symptoms (including fever, stomachache and headache), ear and nose symptoms, breathing issues, and throat symptoms. Two symptom classes (chest pain and anosmia/ageusia), did not load clearly on any of the principal components. Chest pain was therefore excluded from further analyses, while the symptom of anosmia/ageusia, given its relevance to COVID-19 symptomatology, was retained as a single item, separated from the other five symptom classes. The five detected components shared weak to moderate correlations (*r*s between 0.15 and 0.52) and together accounted for 71.05 % of the observed variance.

### Item Response Theory models

The unconstrained GPCM for Asthenia, Influenza, Breathing issues, and Throat symptoms showed the best fit to their relative data, according to AIC, BIC and Log Likelihood. Only in the case of ear/nose symptoms, the fixed-slope (i.e. 1PL) GPCM model showed comparatively better fit to the data and thus was preferred. The comparative statistics of the full and constrained models are presented in **Supplementary Table S2**.

Of the original nine items regarding asthenia, items “vertigo” and “felt dizzy” were removed because of significant misfit to the model. The model estimated on the remaining seven items did not fit the data adequately. The Q3 index showed a high residual correlation – compared to the dataset’s average – between item “been in a bad mood” and “been irritable”, suggesting the presence of a secondary dimension associated with mood alterations. Once these items were removed, leaving a final set of five items, good model-data fit was achieved.

The items regarding influenza-like symptoms did not fit a single GPCM model. Further examination using the DETECT method in package sirt resulted in the presence of a “fever” cluster (including only three items: “feeling feverish”, “sweat” and “chills”), distinct from the remaining 5 items, which assessed more specifically gastro-intestinal issues. We decided to consider these two clusters separately. The Item Response Theory (IRT) model of the “gastro-intestinal” cluster led to dichotomizing item “vomiting” and excluding item “loss of appetite” for misfit, thus resulting in a four-item scale. Due to the very low number of items, we did not fit an IRT model to the fever cluster. Regarding ear and nose symptoms, despite no significant item misfit or problematic category functioning, the initial model did not adequately fit the data. Examination of residual correlations showed high residuals between items “muscle pain” and “joint pain” and between items “painful pressure in the ears” and “painful sinuses”. We therefore excluded items “joint pain” and “painful pressure in the ears” to reduce redundancy. Item “muscle pain” no longer fit the reduced model and was excluded in turn. However, since “muscle or body aches” are indicated as one of the most common symptoms of COVID-19, we decided to retain it as a single item next to the final five items assessing ear and nose symptoms.

Regarding breathing issues, item “difficulty in thinking clearly” and “difficulty in going about your daily business” were removed due to misfit, whereas item “being so unwell you had to stay in bed” was removed due to high residual correlations with item “felt tired”. Item “coughing up mucus” was instead dichotomized to improve the functioning of answer categories. The final model included six items.

All five items associated to throat symptoms fit the model well, and the model offered a good representation of the data.

The final IRT models retained 27 items: 5 assessing asthenia, 3 assessing fever, 4 assessing gastro-intestinal symptoms, 5 assessing nose symptoms, 6 assessing breathing problems and 4 assessing throat symptoms. Items “coughing up mucus” and “vomiting” were recoded as dichotomous (0 = symptom absent. 1 = symptom present).

Items “anosmia/ageusia” and “muscle pain” were not included in any of the models but were kept as single items due to their relevance in diagnosing COVID-19.

**Table 1** shows item parameters and item fit index for the final models. **Table 2** shows model fit indices. Cronbach’s alpha was between 0.69 and 0.92 for all subscales and for the total scale.

**Table 3** reports the questionnaire in its final validated form, which contains only the items retained in the IRT models.

### Questionnaire scores and risk of COVID-19

**Supplementary Table S3** reports descriptive statistics for variables used in the logistic linear regressions. The effects of the total questionnaire score in predicting COVID-19 diagnosis are reported in **Supplementary Table S4**. Results from the model examining individual scale scores (asthenia, gastrointestinal symptoms, fever, ear/nose symptoms, breathing issues, throat symptoms, anosmia/ageusia, and muscle pain) appear in **Supplementary Table S5**.

The total score on the questionnaire was significantly associated with positivity at molecular test for SARS-CoV-2: as expected, individuals displaying a larger number of symptoms or experiencing them more often, as reported in the questionnaire, were significantly more likely to be affected by COVID-19 (exp(β) = 1.203, *p* < .001). Age (being older) and sex (being male) also represented risk factors (exp(β) = 1.038 and 3.057, respectively, *p* < .001). Everything else being equal, none of the comorbidities examined had a significant association with COVID-19 diagnosis.

Investigation of the effects of individual scale scores (**Supplementary Table S5**) revealed that breathing difficulties and neurological symptoms of anosmia/ageusia were significantly associated to higher probabilities of COVID-19 diagnosis, when controlling for other variables in the model. Asthenia, gastrointestinal, fever, Ear/Nose and Throat symptoms, instead, did not have a significant impact, everything else being equal. This means that, in the case of e.g. wheezing, shortness of breath, cough (i.e. breathing issues), loss of smell or taste (i.e. anosmia/ageusia), a greater number or severity of symptoms are associated with an increased chance of testing positive for SARS-CoV-2 swab. The other recorded symptoms, on the other hand, have no impact on the probability of the swab outcome beyond what is already accounted for by the breathing and anosmia/ageusia items. According to this finding, we computed an additional partial score, consisting in the unweighted sum of the breathing symptoms scale and of the anosmia/ageusia item, and used it to estimate scores in association with specific risk thresholds. Results from the model using this partial score as predictor are presented in **Table 4**.

## DISCUSSION

According to the analyses of the present study, six subscales and two individual items, each accounting for a separate dimension of COVID-19 symptomatology, were identified in order to validate the novel questionnaire. Such questionnaire presented some analogy with the ARTIQ^23^, although some clinical categories of the latter, such as “ENT” and the “influenza” symptoms, in our novel assessment model turned into more homogeneous components.

Besides providing a reliable tool for assessing and reporting symptom, the COVID-Q also provided a quantitative score which appeared to correlate with the risk of testing positive to SARS-CoV-2 infection. In particular, higher scores on the “breathing issues” subscale (i.e. “dry cough”, “coughing up mucus”, “problems with breathing”, “wheezing”, “shortness of breath”, “felt tired”) and on the anosmia/ageusia item (i.e. “loss of smell or taste”) were significantly associated with a higher chance of a positive SARS-CoV-2 test.

**Figure 1** shows the average probability of COVID-19 diagnosis as a function of total score in the breathing symptoms scale and in the anosmia/ageusia item, sex and age group. According to our model, a middle-aged person (i.e. 51 years old, the average age in our sample), without any breathing or neurological symptoms had a 0.63% baseline chance of testing positive to a COVID-19 swab, irrespective of gender, smoking behavior or associated illnesses. This baseline chance increased by an average of 1.99 times for every point scored in either the breathing scale or the anosmia/ageusia item. Thus, a score of 1 on the partial subscale represented a 1.24 % chance of a positive swab, which increased to 16.42% for a score of 5 and 99.74% for a score of 16, the highest possible. These risk thresholds were computed for individuals aged 51 years old and did not take into account their sex. However, our model predicted that sex and age would further affect the baseline probability. Namely, the chance of testing positive to a swab increased or decreased by 1.04 times for every year of age over or below 51, respectively. Similarly, males were 2.12 times more likely to be affected by COVID-19, compared to the baseline probabilities presented above, while females were 1.58 times less likely. These results are in line with those reported in the literature, since male gender, for instance, has been associated with worse course of COVID-19, up to higher risk of hospitalization in Intensive Care Unit and even risk of death.^36^ Moreover, regarding age, a similar conclusion has been previouslty obtained, being immunosenescence and inflammaging factors which may determine a less favourable course in older adults.^37^

**Figure 1.**
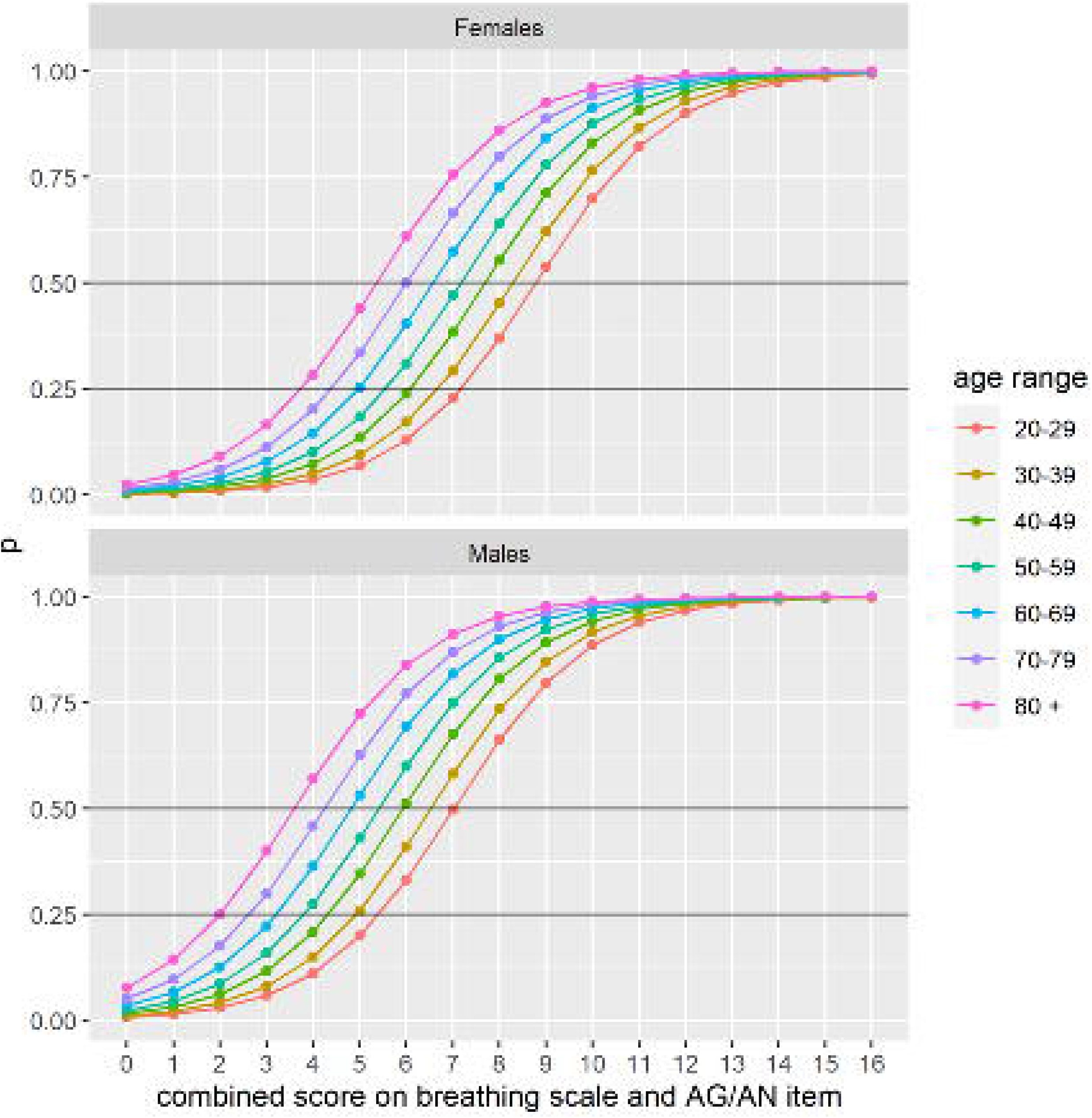
Probability of testing positive to COVID-19 swab (“p” on y axis) as a function of total score on the COVID-Q breathing scale and anosmia/ageusia (AG/AN) item (x axis), age group and sex. Note: probability was computed for the mean age in each age group.

It must be borne in mind that great care must be exercised to ensure an adequate description of the patient’s condition and symptoms, especially in a field, like the emerging COVID-19, where no gold standard exists. The questionnaire presented in this study has been used in previous published articles^13-16^ and allowed to obtain reliable results regarding patients’ symptoms. In those papers, indeed, a proper assessment could be obtained to follow-up a group of patients who had previously tested positive for SARS-CoV-2. Many other available studies on COVID-19 were focused on patient-reported symptoms, also collected via non-conventional means of clinical assessment, including tele-medicine^38^ and the use of social networks^39^. However, to date, a comprehensive and specific tool to collect and scale symptoms from COVID-19 patients has not been described yet.

The COVID-Q may potentially fill this gap, providing a validated and standardized assessment tool to support clinical data collection, also in the absence of a direct contact with the patient, such as in the tele-medicine field.

Although this study also provided score thresholds to define patients at higher risk of testing positive to the SARS-CoV-2 infection, the COVID-Q should not be used as a diagnostic tool. Rather, its application to those who did not receive a microbiological test may help to discriminate COVID-related symptoms, potentially aiming to identify subjects who should be tested. To the best of our knowledge, no validated questionnaires have been developed so far. Usefulness of the questionnaire shown in this paper relies in the high possibility to discriminate the probability of a patient to test positive to SARS-CoV-2. Given the case of a subject who can’t undergo nasopharyngeal swab (e.g., logistical reasons, home bedridden patients), administration of COVID-Q may be a possible and reliable solution for General Practitioners to suggest self-isolation.

The main strengths of this study reside in the controlled design, which allowed to apply the questionnaire to both COVID-19 patients and healthy subjects, and in the accurate definition of its items. The item response theory models led to the development of a panel of questions which allowed to obtain unbiased descriptions of clinical condition, questions focused to meet COVID-19-related symptoms, and a reliable scaling system. On the other hand, it has to be considered that the COVID-Q has been developed and applied in Italian language. Any other language version should be tested to verify whether it retained its measurement properties also in different linguistic and cultural or ethnic backgrounds. Another limitation of the present study is the number of patients. Even if it’s statistically significant, a wider application of the COVID-Q to a larger number of patients may allow to obtain even more precise results and, possibly, to adjust the model we suggested with the present study. It must be observed, also, that being SARS-CoV-2 an RNA virus, it can mutate into diverse variants which may give diverse presentations of COVID-19. Therefore, possible score differences may be encountered by using the questionnaire. At last, the present study has been conducted in a single geographical area, so its application in different areas of other Countries may be useful in order to observe any possible score differences.

In conclusion, in this study we developed and proposed a novel questionnaire to assess and scale the COVID-19-associated symptoms. Its application to COVID-19 patients could improve the reliability in collecting clinical data and assessing the severity of symptoms, while its extension to subjects who have not received a COVID-19 diagnosis yet might help to identify those at higher risk of being affected and, therefore, needing a microbiological test.

## Data Availability

Data are available upon request to the authors.

## Notes

**Conflicts of Interest, Source of Funding and Compliance with Ethical Standards** The authors have no conflict of interest, funding or financial relationships. An informed consent has been obtained for any procedure involving the patients described in this article. The manuscript has not been submitted to more than one journal for simultaneous consideration. The manuscript has not been published previously (party or in full). All of the authors have participated in the planning, writing or revising the manuscript.

### Competing Interest Statement

The authors have declared no competing interest.

### Funding Statement

No funding to disclose.

### Author Declarations

Ethics committee of Treviso and Belluno hospitals.

