## Supplementary tables for "COVID-Q: validation of the first COVID-19 questionnaire based on patient-rated symptom gravity"

### Conflicts of Interest, Source of Funding and Compliance with Ethical Standards

The authors have no conflict of interest, funding or financial relationships. An informed consent has been obtained for any procedure involving the patients described in this article. The manuscript has not been submitted to more than one journal for simultaneous consideration. The manuscript has not been published previously (party or in full). All of the authors have participated in the planning, writing or revising the manuscript.

Table S1 – distribution of comorbidities and risk factors within the sample. For statistical analyses, comorbidities were considered as present or not present.

| | Total (prop) | Control (prop) | Cases (prop) | Fisher p |
|---|---|---|---|---|
| N | 460 | 230 | 230 | |
| Current/ ex smokers | 141 (30.65) | 70 (30.43) | 71 (30.87) | 0.919 |
| Hypertension | 93 (20.22) | 26 (11.3) | 67 (29.13) | 0.000 |
| Diabetes | 26 (5.65) | 5 (2.17) | 21 (9.13) | 0.001 |
| Cardiovascular diseases | 39 (8.48) | 13 (5.65) | 26 (11.3) | 0.030 |
| Cerebrovascular diseases | 7 (1.52) | 2 (0.87) | 5 (2.17) | 0.285 |
| Tumors | 18 (3.91) | 9 (3.91) | 9 (3.91) | 1.000 |
| COPD | 26 (5.65) | 16 (6.96) | 10 (4.35) | 0.313 |
| Renal impairment | 11 (2.39) | 4 (1.74) | 7 (3.04) | 0.544 |
| Liver diseases | 1 (0.22) | 0 (0) | 1 (0.43) | 0.499 |

Note. N = number of participants per group. Prop = proportion. COPD = Chronic Obstructive Pulmonary Disease. The number in brackets indicates the proportion of affected individuals in each sample/subsample.

Table S2: comparative statistics of the full and constrained models.

| Model | AIC | BIC | logLik | X2 | df | p |
|---|---|---|---|---|---|---|
| Asthenia | | | | | | |
| 1PL | 3423.79 | 3502.29 | -1692.90 | | | |
| 2PL | 3409.65 | 3521.19 | -1677.82 | 30.143 | 8 | 0.000 |
| Influenza | | | | | | |
| 1PL | 3928.78 | 4007.27 | -1945.39 | | | |
| 2PL | 3896.02 | 4007.56 | -1921.01 | 48.759 | 8 | 0.000 |
| Ear/ Nose | | | | | | |
| 1PL | 3014.72 | 3084.95 | -1490.36 | | | |
| 2PL | 3017.15 | 3116.30 | -1484.57 | 11.566 | 7 | 0.116 |
| Breathing | | | | | | |
| 1PL | 3887.78 | 3966.27 | -1924.89 | | | |
| 2PL | 3797.78 | 3909.32 | -1871.89 | 105.999 | 8 | 0.000 |
| Throat | | | | | | |
| 1PL | 1262.51 | 1299.69 | -622.25 | | | |
| 2PL | 1236.65 | 1286.23 | -606.33 | 31.857 | 3 | 0.000 |

*Note.* Fit statistics for common-slope (1PL) and unconstrained (2PL) models for each of the 5 components individuated by the PCA.

Table S3. Swab outcome, comorbidities, and clinical manifestations in the enrolled population: descriptive statistics.

| | nbr.val | mean | std.dev | range_2 |
|---|---|---|---|---|
| Swab outcome | 460 | 0,500 | 0,501 | 0 - 1 |
| Age | 460 | 51,031 | 15,206 | 20 - 100 |
| Sex[a] | 460 | 0,378 | 0,485 | 0 - 1 |
| Smoking status[b] | 458 | 0,308 | 0,462 | 0 - 1 |
| Hypertension[c] | 459 | 0,203 | 0,402 | 0 - 1 |
| Diabetes[c] | 459 | 0,057 | 0,231 | 0 - 1 |
| Cardiovascular diseases[c] | 459 | 0,085 | 0,279 | 0 - 1 |
| Cerebrovascular diseases[c] | 459 | 0,015 | 0,123 | 0 - 1 |
| Tumors[c] | 458 | 0,039 | 0,195 | 0 - 1 |
| COPD[c] | 459 | 0,057 | 0,231 | 0 - 1 |
| Renal Impairment[c] | 458 | 0,024 | 0,153 | 0 - 1 |
| Asthenia | 460 | 1,207 | 2,205 | 0 - 10 |
| Gastrointestinal symptms | 460 | 0,946 | 1,552 | 0 - 8 |
| Fever | 460 | 1,007 | 1,682 | 0 - 6 |
| Ear/nose symptoms | 460 | 0,846 | 1,525 | 0 - 9 |
| Breathing symptoms | 460 | 1,674 | 2,356 | 0 - 11 |
| Throat symptoms | 460 | 0,548 | 1,168 | 0 - 7 |
| Anosmia/Ageusia | 460 | 1,213 | 1,870 | 0 - 5 |
| Muscle pain | 460 | 0,370 | 0,675 | 0 - 2 |
| Total score | 460 | 7,809 | 9,588 | 0 - 47 |
| Partial score | 460 | 2,887 | 3,581 | 0 - 16 |

[a] Sex was coded as 0 for females and 1 for males.
[b] Smoking status was coded as 0 for never, and 1 for former or current smokers.
[c] Hypertension, Diabetes, Cardiovascular and Cerebrovascular diseases, Tumors, Chronic Obstructive Pulmonary Disease (COPD) and Renal impairment were coded as 0 for absent and 1 for present.

*Table S4*. Rare-event logistic regression predicting SARS-CoV-2 – COVID-Q total score used as predictor.

| Predictor | *b* | Exp(*b*) | *S.E.* | *Z* | *p* |
|---|---|---|---|---|---|
| (Intercept) | -4.768 | 0.008 | 0.169 | -28.178 | 0.000 |
| Demographics | | | | | |
| Age | 0.037 | 1.038 | 0.011 | 3.456 | 0.001 |
| Sex[a] | 1.118 | 3.057 | 0.275 | 4.059 | 0.000 |
| COVID-19 Symptom questionnaire | | | | | |
| Total score | 0.184 | 1.203 | 0.019 | 9.573 | 0.000 |
| Comorbidities and smoking status | | | | | |
| Smoking status[b] | -0.406 | 0.666 | 0.287 | -1.414 | 0.157 |
| Hypertension[c] | 0.482 | 1.620 | 0.372 | 1.295 | 0.195 |
| Diabetes[c] | 0.296 | 1.344 | 0.698 | 0.424 | 0.672 |
| Cardiovascular diseases[c] | 0.116 | 1.123 | 0.529 | 0.219 | 0.827 |
| Cerebrovascular diseases[c] | -1.386 | 0.250 | 1.193 | -1.162 | 0.245 |
| Tumors[c] | -0.735 | 0.479 | 0.620 | -1.186 | 0.236 |
| COPD[c] | -1.045 | 0.352 | 0.656 | -1.592 | 0.111 |
| Renal Impairment[c] | 0.097 | 1.102 | 1.102 | 0.088 | 0.930 |

*Note*. COPD = chronic obstructive pulmonary disease

[a] Gender was coded as 0 for females and 1 for males.

[b] Smoking status was coded as 0 for never, and 1 for former or current smokers.

[c] Hypertension, Diabetes, Cardiovascular and Cerebrovascular diseases, Tumors, COPD and Renal impairment were coded as 0 for absent and 1 for present.

*Table S5*. Rare-event logistic regression predicting SARS-CoV-2 infection. COVID-Q's subscale scores used as predictors.

| Predictor | *b* | Exp(*b*) | *S.E.* | *Z* | *p* |
|---|---|---|---|---|---|
| (Intercept) | -3.194 | 0.041 | 0.178 | -17.989 | 0.000 |
| Demographics | | | | | |
| Age | 0.041 | 1.041 | 0.012 | 3.267 | 0.001 |
| Sex[a] | 1.144 | 3.140 | 0.323 | 3.543 | 0.000 |
| COVID-Q | | | | | |
| Asthenia | -0.098 | 0.907 | 0.109 | -0.904 | 0.366 |
| Gastrointestinal symptoms | 0.008 | 1.008 | 0.149 | 0.055 | 0.956 |
| Fever | 0.289 | 1.334 | 0.151 | 1.908 | 0.056 |
| Ear/nose symptoms | 0.086 | 1.089 | 0.135 | 0.636 | 0.525 |
| Breathing symptoms | 0.553 | 1.738 | 0.132 | 4.184 | 0.000 |
| Throat symptoms | -0.249 | 0.780 | 0.187 | -1.329 | 0.184 |
| Anosmia/Ageusia | 0.807 | 2.240 | 0.124 | 6.524 | 0.000 |
| Muscle pain | -0.311 | 0.733 | 0.347 | -0.896 | 0.370 |
| Comorbidities and smoking status | | | | | |
| Smoking status[b] | -0.410 | 0.664 | 0.335 | -1.225 | 0.221 |
| Hypertension[c] | 0.411 | 1.508 | 0.423 | 0.971 | 0.331 |
| Diabetes[c] | 0.438 | 1.549 | 0.839 | 0.522 | 0.602 |
| Cardiovascular diseases[c] | -0.005 | 0.995 | 0.620 | -0.007 | 0.994 |
| Cerebrovascular diseases[c] | -1.597 | 0.202 | 1.513 | -1.056 | 0.291 |
| Tumors | -1.152 | 0.316 | 0.737 | -1.563 | 0.118 |
| COPD | -1.243 | 0.288 | 0.751 | -1.655 | 0.098 |
| Renal Impairment | 0.314 | 1.369 | 1.328 | 0.237 | 0.813 |

*Note*. COPD = chronic obstructive pulmonary disease

[a] Gender was coded as 0 for females and 1 for males.

[b] Smoking status was coded as 0 for never, and 1 for former or current smokers.

[c] Hypertension, Diabetes, Cardiovascular and Cerebrovascular diseases, Tumors, COPD and Renal impairment were coded as 0 for absent and 1 for present.
