## Supplementary material for "COVID-Q: validation of the first COVID-19 questionnaire based on patient-rated symptom gravity": Tables

### Conflicts of Interest, Source of Funding and Compliance with Ethical Standards

The authors have no conflict of interest, funding or financial relationships. An informed consent has been obtained for any procedure involving the patients described in this article. The manuscript has not been submitted to more than one journal for simultaneous consideration. The manuscript has not been published previously (party or in full). All of the authors have participated in the planning, writing or revising the manuscript.

**TABLES**

Table 1. Item parameters and fit from final Item Response Theory (IRT) models.

| | Discrimination | Threshold 1 | Threshold 2 | S_X2 | p |
|---|---|---|---|---|---|
| *Asthenia* | | | | | |
| being awake most of the night | 5.600 | 1.370 | 1.794 | 5.008 | 0.286 |
| difficulty falling asleep | 3.657 | 1.262 | 1.849 | 3.511 | 0.476 |
| waking up several times at night | 4.568 | 1.028 | 1.700 | 3.903 | 0.419 |
| poor quality of sleeping | 3.904 | 1.119 | 1.626 | 5.492 | 0.359 |
| not feeling yourself | 1.320 | 1.067 | 1.122 | 7.446 | 0.489 |
| *Gastrointestinal symptoms* | | | | | |
| vomit[1] | 4.203 | 1.805 | - | 4.849 | 0.183 |
| nausea | 2.703 | 1.485 | 1.905 | 9.807 | 0.133 |
| diarrhoea | 1.915 | 0.952 | 1.885 | 8.865 | 0.181 |
| abdominal pain | 2.340 | 1.724 | 2.502 | 1.373 | 0.712 |
| *Ear and nose symptoms* | | | | | |
| headache | 1.254 | 1.417 | 1.455 | 5.923 | 0.549 |
| runny nose | 2.136 | 1.420 | 2.030 | 2.634 | 0.621 |
| blocked nose | 2.136 | 1.101 | 2.026 | 4.287 | 0.509 |
| sneezing | 2.136 | 1.252 | 2.492 | 8.535 | 0.129 |
| painful pressure in ears | 2.136 | 1.919 | 2.627 | 6.381 | 0.094 |
| watery eyes | 2.136 | 1.765 | 3.082 | 3.696 | 0.158 |
| *Breathing issues* | | | | | |
| problems with breathing | 5.022 | 1.083 | 1.731 | 5.001 | 0.544 |
| wheezing | 6.659 | 1.193 | 1.829 | 7.909 | 0.161 |
| shortness of breath | 9.601 | 0.896 | 1.700 | 5.696 | 0.127 |
| coughing up mucus[1] | 1.333 | 1.910 | - | 2.640 | 0.955 |
| dry cough | 1.379 | 1.032 | 1.693 | 13.527 | 0.260 |
| felt tired | 1.683 | 0.547 | 0.980 | 12.926 | 0.114 |
| *Throat symptoms* | | | | | |
| swollen glands | 1.691 | 3.038 | 3.300 | 0.149 | 0.699 |
| sore throat | 3.870 | 1.313 | 1.820 | 3.711 | 0.054 |
| hoarseness | 1.154 | 2.038 | 2.678 | 8.197 | 0.085 |
| tickles in the throat | 5.046 | 1.041 | 1.893 | 1.195 | 0.274 |

*Note.* Discrimination = ability of the item to discriminate between individuals with different symptom frequencies. Threshold 1 = symptom frequency value that marks the cutoff between adjacent answer categories: 0 = “no symptom” and 1 = “I experience this symptom a little.” Threshold 2 = cutoff value between category 1 = “I experience this symptom a little” and 2 = “I experience this symptom a lot”

[1] Dichotomous item: 0 = symptom absent, 1 = symptom present. Threshold 2 was not computed for dichotomized items.

Table 2. Fit indices for the final Item Response Theory (IRT) models

| | M2* | df | p | RMSEA | SRMSR | TLI | CFI |
|---|---|---|---|---|---|---|---|
| Asthenia | 2,028* | 5 | 0.845 | 0.000 | 0.016 | 1.003 | 1.000 |
| Stomachache | 3,664 | 1 | 0.056 | 0.076 | 0.084 | 0.955 | 0.993 |
| Nose Symptoms | 8,478 | 4 | 0.076 | 0.049 | 0.093 | 0.967 | 0.974 |
| Breathing issues | 4,669 | 4 | 0.323 | 0.019 | 0.075 | 0.998 | 0.999 |
| Throat symptoms | 3,334* | 2 | 0.189 | 0.038 | 0.042 | 0.991 | 0.997 |

*Note.* p values > .05 indicate good model-data fit.

* The model did not have sufficient degrees of freedom for the computation of the M2* index. The less restrictive C2 index was used instead. The interpretation of p values remains unvaried[34].

Table 3 The final COVID-Q questionnaire.

| *Asthenia* | | | |
|---|---|---|---|
| "Being awake most of the night" | 0 (none) | 1 (a little) | 2 (a lot) |
| "Difficulty falling asleep" | 0 (none) | 1 (a little) | 2 (a lot) |
| "Waking up several times at night" | 0 (none) | 1 (a little) | 2 (a lot) |
| "Poor quality of sleeping" | 0 (none) | 1 (a little) | 2 (a lot) |
| "Not feeling yourself" | 0 (none) | 1 (a little) | 2 (a lot) |
| *Gastrointestinal symptoms* | | | |
| "vomit" | 0 (absent) | 1 (present) | |
| "nausea" | 0 (none) | 1 (a little) | 2 (a lot) |
| "diarrhea" | 0 (none) | 1 (a little) | 2 (a lot) |
| "abdominal pain" | 0 (none) | 1 (a little) | 2 (a lot) |
| *Fever* | | | |
| "Feeling feverish" | 0 (none) | 1 (a little) | 2 (a lot) |
| "Sweat" | 0 (none) | 1 (a little) | 2 (a lot) |
| "Chills" | 0 (none) | 1 (a little) | 2 (a lot) |
| *Ear and nose symptoms* | | | |
| "headache" | 0 (none) | 1 (a little) | 2 (a lot) |
| "runny nose" | 0 (none) | 1 (a little) | 2 (a lot) |
| "blocked nose" | 0 (none) | 1 (a little) | 2 (a lot) |
| "sneezing" | 0 (none) | 1 (a little) | 2 (a lot) |
| "watery eyes" | 0 (none) | 1 (a little) | 2 (a lot) |
| *Breathing issues* | | | |
| "problems with breathing" | 0 (none) | 1 (a little) | 2 (a lot) |
| "wheezing" | 0 (none) | 1 (a little) | 2 (a lot) |
| "shortness of breath" | 0 (none) | 1 (a little) | 2 (a lot) |
| "coughing up mucus" | 0 (absent) | 1 (present) | |
| "dry cough" | 0 (none) | 1 (a little) | 2 (a lot) |
| "felt tired" | 0 | 1 | 2 |

| | | | | | | |
|---|---|---|---|---|---|---|
| | (none) | (a little) | (a lot) | | | |
| *Throat symptoms* | | | | | | |
| "swollen glands" | 0<br>(none) | 1<br>(a little) | 2<br>(a lot) | | | |
| "sore throat" | 0<br>(none) | 1<br>(a little) | 2<br>(a lot) | | | |
| "hoarseness" | 0<br>(none) | 1<br>(a little) | 2<br>(a lot) | | | |
| "tickles in the throat" | 0<br>(none) | 1<br>(a little) | 2<br>(a lot) | | | |
| *Muscle pain* | 0<br>(none) | 1<br>(a little) | 2<br>(a lot) | | | |
| *Anosmia/Ageusia* | *0*<br>(no symptom) | *1* | *2* | *3* | *4* | *5*<br>*(complete absence of smell or taste)* |

*Table 4*. Rare-event logistic regression predicting SARS-CoV-2 infection with unweighted sum of the breathing symptoms scale and of the anosmia/ageusia item. COVID-19 symptom questionnaire's partial score used as predictor.

| Predictor | *b* | Exp(*b*) | *S.E.* | *Z* | *p* |
|---|---|---|---|---|---|
| (Intercept) | -5.066 | 0.006 | 0.185 | -27.451 | 0.000 |
| Demographics | | | | | |
| Age[d] | 0.039 | 1.040 | 0.012 | 3.272 | 0.001 |
| Sex[a,d] | 1.211 | 3.357 | 0.309 | 3.915 | 0.000 |
| COVID-Q | | | | | |
| Partial score | 0.688 | 1.990 | 0.070 | 9.861 | 0.000 |
| Comorbidities and smoking status | | | | | |
| Smoking status[b,d] | -0.526 | 0.591 | 0.328 | -1.602 | 0.109 |
| Hypertension[c,d] | 0.521 | 1.683 | 0.414 | 1.258 | 0.208 |
| Diabetes[c,d] | 0.579 | 1.784 | 0.811 | 0.714 | 0.475 |
| Cardiovascular diseases[c,d] | -0.034 | 0.966 | 0.590 | -0.058 | 0.953 |
| Cerebrovascular diseases[c,d] | -1.652 | 0.192 | 1.401 | -1.179 | 0.238 |
| Tumors[c,d] | -0.894 | 0.409 | 0.691 | -1.294 | 0.196 |
| COPD[c,d] | -1.573 | 0.207 | 0.756 | -2.082 | 0.037 |
| Renal Impairment[c,d] | 0.736 | 2.087 | 1.210 | 0.608 | 0.543 |

*Note*. COPD = chronic obstructive pulmonary disease

[a] Gender was coded as 0 for females and 1 for males. Reference was made for male gender.

[b] Smoking status was coded as 0 for never, and 1 for former or current smokers.

[c] Hypertension, Diabetes, Cardiovascular and Cerebrovascular diseases, Tumors, COPD and Renal impairment were coded as 0 for absent and 1 for present.

[d] These predictors were mean centered for the regression analysis.
